# High parental vaccine motivation at a neighborhood-based vaccine and testing site serving a predominantly Latinx community

**DOI:** 10.1101/2021.07.30.21261274

**Authors:** Jamie Naso, Susy Rojas, James Peng, Carina Marquez, Maria Contreras, Edgar Castellanos, Susana Rojas, Luis Rubio, Diane Jones, Jon Jacobo, Douglas Black, Valerie Tulier-Laiwa, Jacqueline Martinez, Gabriel Chamie, Genay Pilarowski, Joseph DeRisi, Diane Havlir, Maya Petersen

## Abstract

**Purpose:** To understand vaccine attitudes of Latinx parents highly impacted by COVID-19. *Methods*. In April 2021, we surveyed parents about their attitudes for COVID-19 vaccination of their children at a community-based outdoor testing/vaccination site serving predominantly low-income, Latinx persons in San Francisco.

**Results:** Among 1,033 parents (75% Latinx), 92% would “definitely” or “probably” vaccinate their children. Vaccine hesitancy was higher for younger children; concerns included side effects and impacts on fertility. Doctors and community organizations were noted as trusted sources of information, including among vaccine-hesitant parents.

**Conclusion:** Latinx parents accessing neighborhood-based COVID-19 testing/vaccination services are highly motivated to vaccinate their children for COVID-19.

## Introduction

In California, Latinx persons have been disproportionately affected by the COVID-19 pandemic,^1^ due to structural inequalities such as having frontline occupations with increased risk of exposure unaffected by stay-at-home orders.^2^ Neighborhood-based testing and vaccination sites that overcome structural barriers to access and that foster trust through culturally competent community staffing have proven to be a very effective means to rapidly achieve high vaccine uptake for adults in highly impacted Latinx neighborhoods^3–5^ and could be leveraged to reach children as vaccines are determined to be safe and effective for younger age groups.^6,7^ We sought to characterize parental attitudes about vaccinating children at a community-based testing/vaccination outdoor site serving a predominantly Latinx population in San Francisco.

## Methods

### Study Setting and Population

Unidos en Salud, a community (Latino Task Force), academic (UCSF, UC Berkeley, Chan-Zuckerberg Biohub), and San Francisco Department of Public Health partnership, has provided low-barrier, culturally appropriate, community-based SARS-Cov-2 test and respond services in San Francisco’s Mission District since April 2020, and vaccinations since January 2021, with tailored outreach to the Latinx community.^2,8^ On May 16, we began offering vaccines to youth age 12-15. Over 22,000 vaccine doses have been administered (adults and youth; 72% Latinx, 61% household income <$50,000) at the site as of June 4, 2021.

From April 4-28, 2021, all adults (aged ≥18) seeking free BinaxNOW rapid COVID-19 testing^9,10^ or who had just received either the first or second dose of a vaccine shot (Pfizer-BioNTech) were asked if they had children. Those reporting any children aged<18 years were offered a survey on their attitudes regarding COVID-19 vaccinations for their children.

### Measures

Participants completed the surveys in English or Spanish; bilingual staff were available for assistance. The survey asked parents with at least one child (<18 years) questions on COVID-19 vaccine acceptability for their children of each age group (ages 16-17, 12-15, 5-11, 0-4 years). Predictors of vaccine hesitancy among parents were evaluated using multivariate regression, fit using generalized estimating equations with a log-link function and cluster-robust standard errors (to allow for multiple children per parent), with age group, parent gender, and ethnicity included as independent variables.

### Human Subjects

The study was conducted under a public health surveillance program reviewed by UCSF Committee on Human Research. Survey participants provided consent in their preferred language.

## Results

Of the 1,966 parents with a child <18 years old, 1,033 (53%) completed the survey. Completion rates were slightly lower among men (49%) versus women (56%) and among Latinx parents (51%) versus parents of other ethnicities (60%). Among those surveyed, 875 (85%) completed the survey immediately post-vaccination and 158 (15%) completed prior to testing. Seven hundred eighty eight respondents (76%) identified as Latinx, 88 (9%) had a child who had previously contracted COVID-19 (Table 1), 291 (28%) lived in the zip code immediately surrounding the site, and 814 (79%) lived in San Francisco (Figure 1).

**Table 1:** Characteristics of parents who completed the survey (N = 1,033)

| **Characteristic** |  |
| --- | --- |
| Gender (%) |  |
| Female | 534 (51.7%) |
| Male | 492 (47.6%) |
| Other/Unknown | 7 (0.678%) |
| Ethnicity (%) |  |
| Latinx | 788 (76.3%) |
| White | 124 (12.0%) |
| Asian | 53 (5.13%) |
| Black | 16 (1.55%) |
| Other | 52 (5.03%) |
| Median Age (IQR) | 41 (35-46) |
| Has child in age group (%) |  |
| Age 16-17 | 211 (20.4%) |
| Age 12-15 | 381 (36.9%) |
| Age 5-11 | 513 (49.7%) |
| Age 0-4 | 353 (34.2%) |
| Residence (%) |  |
| In neighborhood zip code 94110 | 291 (28.2%) |
| Outside 94110 but in San Francisco | 523 (50.6%) |
| Outside San Francisco | 219 (21.2%) |
| Recruitment (%) |  |
| At testing site | 158 (15.3%) |
| At vaccine site | 875 (84.7%) |
| Received at least one dose of vaccine (%) | 983 (95.2%) |
| Child has gotten COVID (%) | 88 (8.5%) |

**Figure 1:**
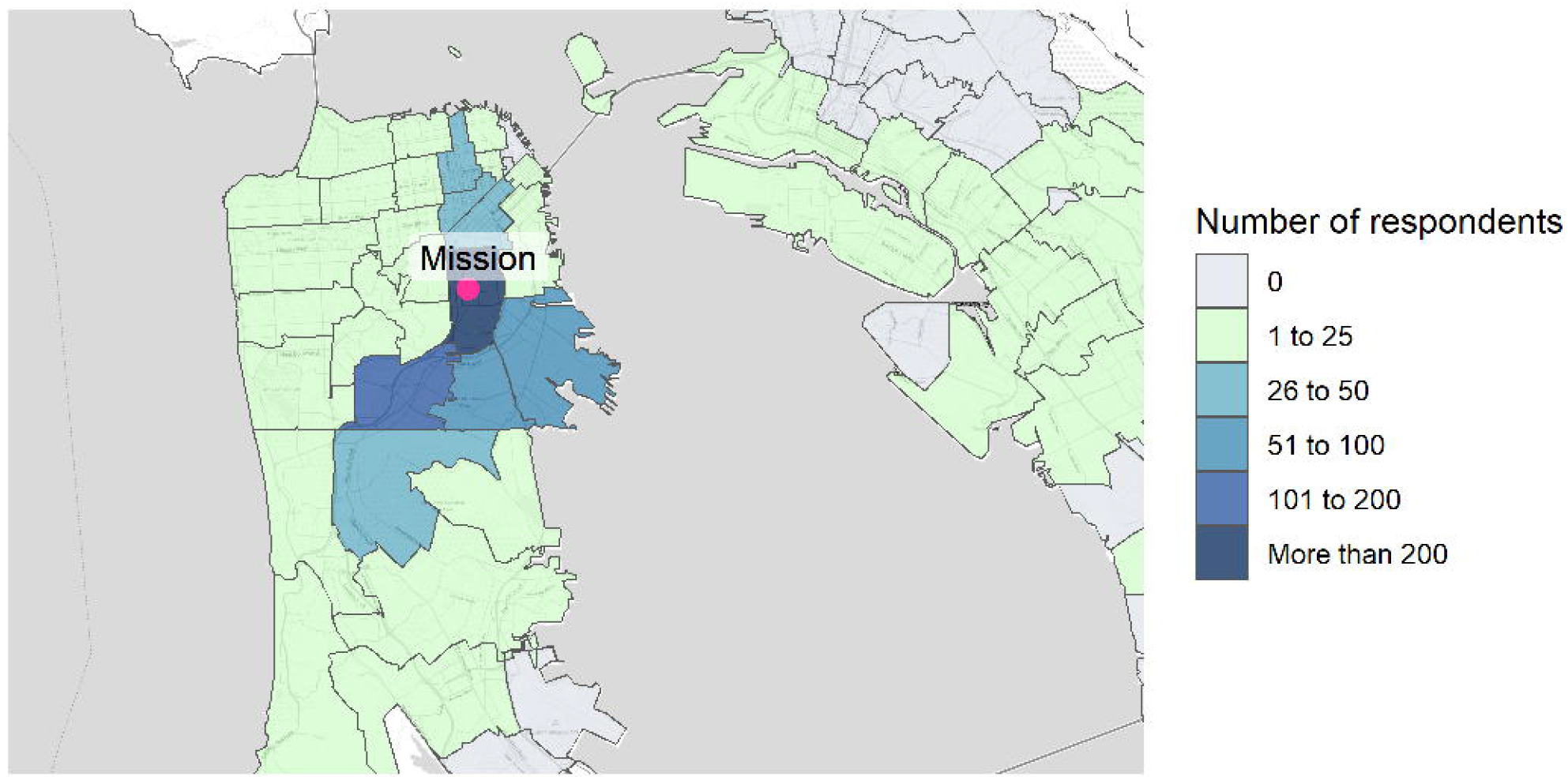
Map of those who completed the survey. Map tiles by Stamen Design, under CC BY 3.0. Data by OpenStreetMap, under CC BY SA.

Vaccine motivation among parents (defined as “definitely” or “probably” intending to vaccinate their child) was 90% (312/348), 91% (461/506), 93% (348/375), and 97% (197/203) for parents with children aged 0-4, 5-11, 12-15, and 16-17, respectively (Figure 2). Among vaccine-motivated parents with children aged 12-15 and 16-17, 65% (220/336) and 83% (158/190), respectively indicated that they would seek an approved vaccine for their child “as soon as they could”. The proportion of parents motivated to seek immediate vaccination for their children was lower for parents with younger children; 47% (144/304) of vaccine-motivated parents with children aged 0-4 and 53% (237/446) of parents with children aged 5-11 (Figure 3). Overall, 92% (949/1,033) of parents of children of all ages were vaccine-motivated, and 58% (546/949) indicated that they would vaccinate their children as soon as possible. Motivations for vaccine-seeking parents included keeping their child safe (92%, 874/949) protecting the community (66%, 629/949) protecting the adults in their family (64%, 605/949), and vaccination potentially being required for school or childcare (61%, 583/949).

**Figure 2:**
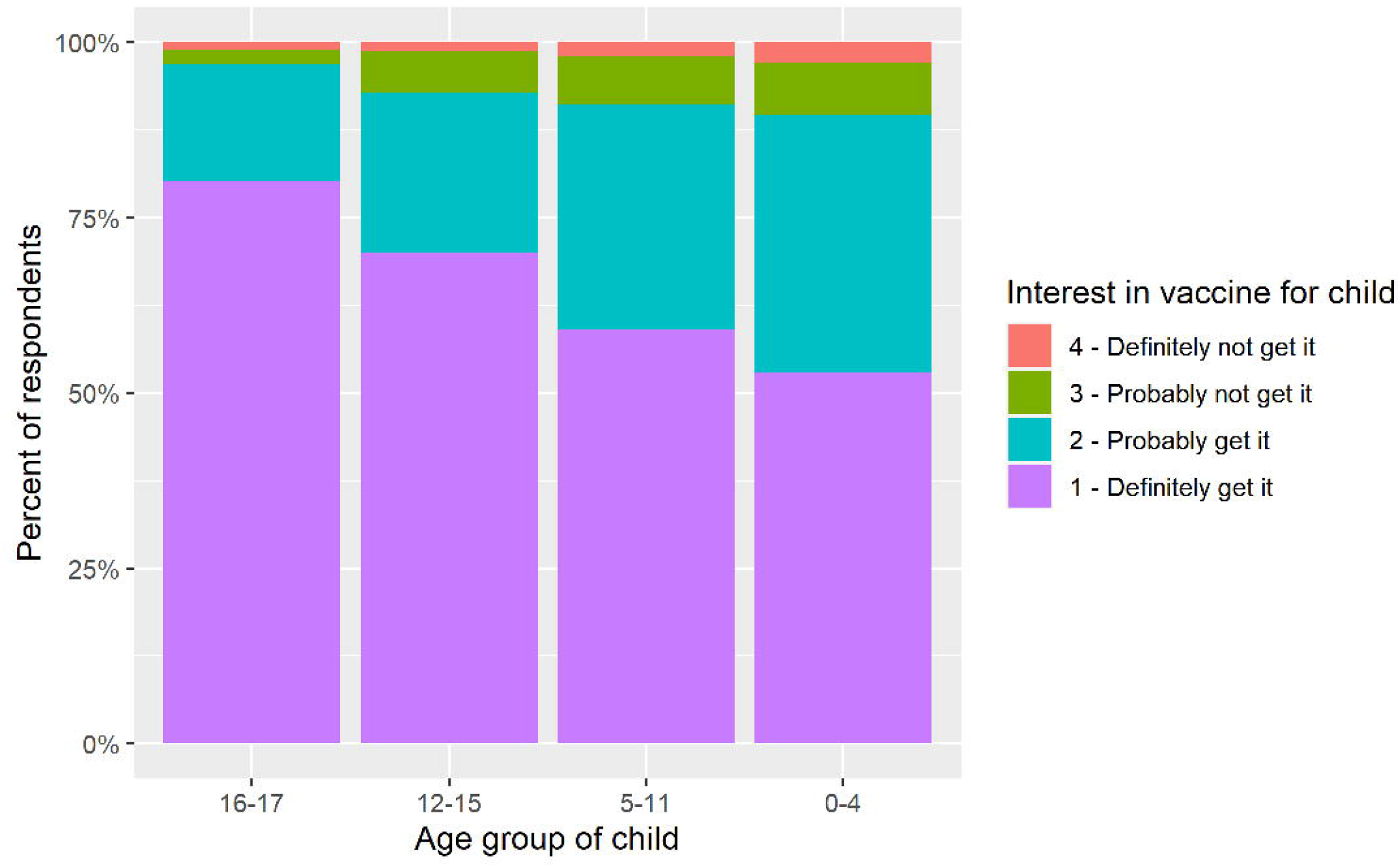
Vaccine interest among parents with children in each age group (16-17, 12-15, 5-11, and 0-4 years)

**Figure 3:**
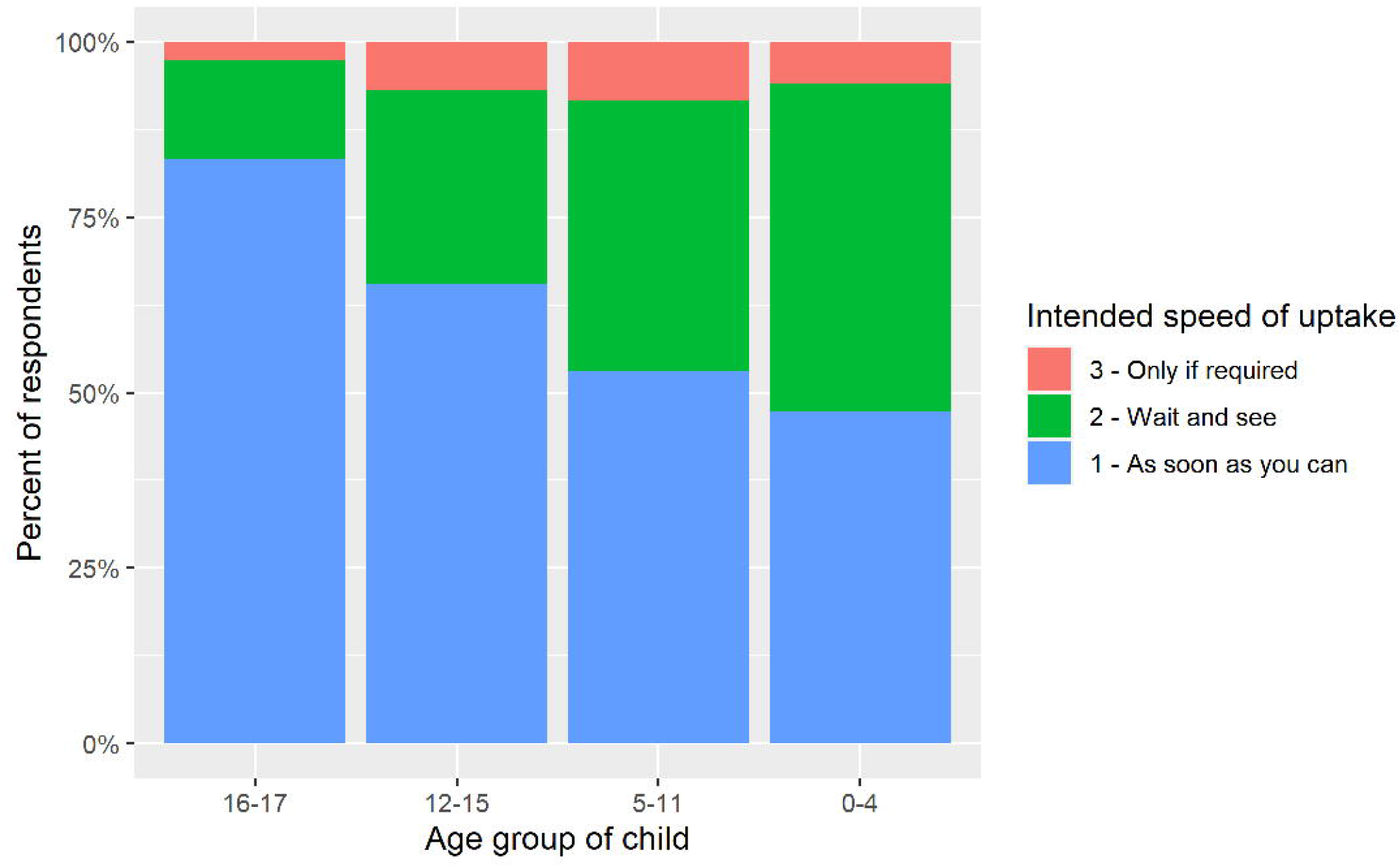
Intended speed of vaccine uptake for children among vaccine motivated parents

Among vaccine-hesitant parents, concerns included immediate side effects (60%, 55/91), long-term effects (41%, 37/91), fear of the vaccine affecting the child’s fertility (19%, 17/91), and belief that the child is not at risk for severe disease (8%, 7/91). Vaccine-hesitant parents had lower amounts of trust in all information sources than vaccine-motivated respondents, although 80% of them indicated a “great” or “good” deal of trust for their children’s doctors and 50% indicated trust for community-based groups (Table 2).

**Table 2:** Respondents who indicated a “good” or “great” deal of trust in different information sources.*

| Group | Vaccine-hesitant for all age groups of children | Vaccine motivated for at least one age group of child | Overall | p-value |
| --- | --- | --- | --- | --- |
| Child’s doctor | 80.3% (57/71) | 93.3% (805/863) | 92.3% (862/934) | <0.001 |
| Latino Task Force (or other community organization) | 51.7% (31/60) | 79.3% (605/763) | 77.3% (636/823) | <0.001 |
| State government | 49.2% (31/63) | 78.7% (631/802) | 76.5% (662/865) | <0.001 |
| Federal government | 34.9% (22/63) | 77.0% (607/788) | 73.9% (629/851) | <0.001 |
| Friends and family | 50.0% (32/64) | 71.5% (580/811) | 69.9% (612/875) | <0.001 |
| Newspaper, tv, and radio | 32.8% (21/64) | 49.3% (388/787) | 48.1% (409/851) | 0.016 |
| Social media | 15.4% (10/65) | 25.8% (201/780) | 25.0% (211/845) | 0.087 |

\* Denominators exclude those who skipped each respective question.

Vaccine acceptance was similar among Latinx (92%, 723/788) and non-Latinx (92%, 226/245) parents, and among men (93%, 457/492) and women (91%, 485/534). In multivariate analysis, having children in younger age groups (child age 12-15 aOR: 2.41, 95%CI:1.01-5.74; age 5-11 aOR: 2.97, 95%CI:1.29-6.85; age 0-4 aOR: 3.43, 95%CI:1.46-8.09; ref. child age 16-17) was associated with increased hesitancy. Female gender (aOR: 1.28, 95%CI:0.90-1.83) and non-Latinx ethnicity (aOR: 1.15, 95%CI:0.76-1.74) were not significantly associated with parental vaccine hesitancy in this population. Open text comments expressed concern for short and longer term side effects, including impacts on fertility (Table 3).

**Table 3:** Selected open text quotes from those surveyed

| Comments among vaccine motivated | Comments among vaccine hesitant |
| --- | --- |
| I am not ready to enroll my child in a vaccine study but eager to get her vaccinated when it's been studied.<br>Naturally, we are slightly more cautious with vaccinating our children than with ourselves, especially since at this point there are already millions of other adults who have been vaccinated without known side effects. But in the end, once the scientific community says it is safe, we will certainly proceed.<br>If the vaccine has harsh side effects for adults such as fever, sore arm and so on, how will it be for children to withstand that?<br>[I] would like to see how effective it is on the first 50 to 100 thousand children before giving to ok for my daughter to be vaccinated.<br>*(Translated from Spanish)* Thank you very much I am grateful to God and to you all for saving so many lives.<br>I appreciate the work of the Latino Task force, the volunteers, medical workers who are on the front lines. Thank you for all your hard work!<br>*(Translated from Spanish)* I want to get it, but I want to know for sure that it will not have any bad effect on my daughter. She is only 12 years old and all I want is to be sure that it [vaccine] will not affect her health. | Children should not get vaccinated and the government should not force them to do so. It's a new drug and it is not known when, how, or what will be affected. It should be a person's choice once they turn 18.<br>*(Translated from Spanish)* I am worried about reactions to the vaccine in small children and I am afraid because it is a new virus and we don't know the [virus'] future effects.<br>What are the effects on fertility for our son?<br>*(Translated from Spanish)* I would like for my children to be vaccinated but only once the vaccine is safe for them and has been effective for more than 2 years. At the moment no [to vaccination].<br>*(Translated from Spanish)* I am worried that the vaccine will cause harm to younger children. I have heard that younger children have been vaccinated who should not have been [vaccinated]. |

## Discussion

Among a population of predominantly Latinx parents attending a community-based testing/vaccination site in a neighborhood highly impacted by COVID-19, over 90% of parents were probable or likely to vaccinate their children. Parents were motivated to protect their children and the community. Primary concerns focused on safety and fertility, and acceptance was lower for younger children. Trusted sources of information included medical providers and community groups.

Hesitancy of U.S. Latino parents to vaccinate their children ranges from 26-47% in surveys conducted via phone, Facebook, and online.^11–13^ We were specifically interested in understanding attitudes of Latinx parents highly affected by the pandemic, many of whom were monolingual and do not have a primary care provider. It is difficult to directly compare findings of these reports to our results because they differ in their intent, sampling approaches, and survey methods. In this setting of a community-based testing/vaccination site, building on long-term community engagement and established trust, we found both Latinx and non-Latinx parents were highly motivated to vaccinate their children, expressing low rates of vaccine hesitancy overall (8%).

Parental concerns included short and long-term side effects, including potential fertility effects, similar to previous reports.^11^ Parents of younger children were more likely to be vaccine hesitant; the lack of data on vaccine effectiveness and safety in younger children likely contributed. Responding effectively to parental concerns will require acknowledging their grounding in historical experience, including the sterilization of Latina women without consent during the 1970s^14^ and more recently, allegations of forced hysterectomies at U.S. Immigration and Custom Enforcement detention centers.^15^ Understanding and leveraging trusted sources of information will be crucial as new vaccine safety reports become available, such as those reporting myocarditis in youth.^16,17^

Both pediatricians and community-based organizations were trusted sources of information, including among vaccine hesitant parents. Many clients at our community sites report having no primary physician.^3,18^ For these reasons, bilingual physicians and health providers are present at our community vaccination site, and we provide health system registration. Our community partner (Unidos en Salud) organization’s staff receive regular updates on COVID vaccines, reviewing questions asked by participants at the site and concerns brought up by those in the neighborhood.

Optimizing vaccine uptake among children, as for adults, should include multiple options. Data from other surveys conducted among clients attending the same low-barrier vaccine site preferred this neighborhood site over schools for vaccination of their children.^3,18^ Our community vaccination site offered onsite registration (eliminating needs for parental computer literacy), bilingual staff (overcome language barriers), weekend and weekday options for single and working parents, and a culturally sensitive and community-facing approach with key input from community leaders and stakeholders. These preferences emphasize how low-barrier vaccine site options are important additions to established healthcare settings, school-based and mobile vaccination options.

After the completion of our survey, on May 16, we began offering vaccinations to youth age 12-15 years with the EUA-granted Pfizer-BioNTech vaccine. Among the 320 youth vaccinated as of June 4 (73% Latinx), 74% were from a household who either had been tested or vaccinated at our site. Investment in neighborhood based, low-barrier sites provided the opportunity for families to seek information and vaccination services locally for their children at a location that had garnered their trust.

Our study was subject to limitations. First, the survey was intentionally conducted among parents the vast majority of whom had themselves received a vaccine. Findings reported should thus not be extrapolated to the general population, but rather illustrate the potential to leverage community-based sites to effectively reach children in highly impacted areas for vaccination. Second, both scientific context and community attitudes can evolve rapidly. Rather than providing a single answer expected to remain static over time and place, the current study illustrates the potential to leverage community vaccination sites to rapidly generate actionable and locally and culturally relevant context-specific knowledge. Finally, we did not include surveys of the youth themselves, who have an important perspective that may influence vaccine uptake.^19^

In conclusion, Latinx parents disproportionately affected by COVID-19 and utilizing a neighborhood testing/vaccination site were highly motivated to vaccinate their children to protect their health and that of the community.

## Data Availability

Due to the nature of this research, participants of this study did not agree for their data to be shared publicly, so supporting data is not available.

## Acknowledgments

We would like to thank the community members who participated in this initiative and the many vaccination and testing site staff, community ambassadors, and volunteers. We thank the Chan Zuckerberg Initiative, Supervisor Hillary Ronen, Mayor London Breed, The San Francisco Department of Public Health and Dr. Grant Colfax, Dr. Naveena Bobba, Dr. Susan Phillips, Dr. Mary Mercer, Emily Reingold, and Dr. Ellen Chen. We gratefully acknowledge the San Francisco Latino Task Force-Response to COVID-19, Bay Area Phlebotomy and Laboratory Services (BayPLS), Stacy Powers and BRAVA for Women in the Arts, Bevan Dufty and the BART team, PrimaryBio COVID-19 Testing Platform, Dr. Jessica Briggs and Dr. Susa Coffey from UCSF, and Dara Fonseca, Jocelin Payan, and Jack Fukushima from Unidos en Salud.

This work was supported by University of California, San Francisco, the Chan Zuckerberg Initiative, and the San Francisco Department of Public Health. The funders had no role in study design, data collection and analysis, decision to publish, or preparation of the manuscript.

